# Proteomic and Metabolomic Investigation of COVID-19 Patients with Elevated Serum Lactate Dehydrogenase

**DOI:** 10.1101/2021.01.10.21249333

**Authors:** Haixi Yan, Xiao Liang, Juping Du, Zebao He, Yu Wang, Mengge Lyu, Liang Yue, Fangfei Zhang, Zhangzhi Xue, Luang Xu, Guan Ruan, Jun Li, Hongguo Zhu, Jiaqin Xu, Shiyong Chen, Chao Zhang, Dongqing Lv, Zongmei Lin, Bo Shen, Yi Zhu, Biyun Qian, Haixiao Chen, Tiannan Guo

## Abstract

Serum lactate dehydrogenase (LDH) has been established as a prognostic indicator given its differential expression in COVID-19 patients. However, the molecular mechanisms underneath remain poorly understood. In this study, 144 COVID-19 patients were enrolled to monitor the clinical and laboratory parameters over three weeks. Serum lactate dehydrogenase (LDH) was shown elevated in the COVID-19 patients on admission and declined throughout disease course, and its ability to classify patient severity outperformed other biochemical indicators. A threshold of 247 U/L serum LDH on admission was determined for severity prognosis. Next, we classified a subset of 14 patients into high- and low-risk groups based on serum LDH expression and compared their quantitative serum proteomic and metabolomic differences. The results found COVID-19 patients with high serum LDH exhibited differentially expressed blood coagulation and immune responses including acute inflammatory responses, platelet degranulation, complement cascade, as well as multiple different metabolic responses including lipid metabolism, protein ubiquitination and pyruvate fermentation. Specifically, activation of hypoxia responses was highlighted in patients with high LDH expressions. Taken together, our data showed that serum LDH levels are associated COVID-19 severity, and that elevated serum LDH might be consequences of hypoxia and tissue injuries induced by inflammation.

## INTRODUCTION

COVID-19 is an ongoing global pandemic caused by severe acute respiratory syndrome coronavirus 2 (SARS-CoV-2). A high viral transmission rate and the lack of effective therapy contributed to more than 83 million infected cases as of time January 3^rd^, 2021 ^[1]^.

To better diagnose COVID-19 and monitor the disease progress, multiple molecules have been proposed as prognostic indicators ^[2]^. Lactate dehydrogenase (LDH) is an intracellular enzyme, catalyzing pyruvate fermentation and facilitating glycolysis. LDH is released into the blood after cell death and has been reported to increase in a variety of diseases including Severe acute respiratory syndrome (SARS) ^[3]^, diabetes ^[4]^, and cancers ^[5]^. Serum LDH levels in COVID-19 patients are over-expressed ^[2]^, especially in severe and critical patients ^[6-9]^. They decrease throughout disease course ^[7,10]^, in correlation with viral mRNA clearance ^[7]^. Related studies have shown that serum LDH is well correlated with respiratory failure ^[11]^, lung injury, disease severity ^[12]^ and mortality ^[13]^ in COVID-19 patients.

However, the molecular mechanisms underlying the LDH’s association with the COVID-19 disease progression remains poorly understood. Most studies attribute serum LDH elevation to its release from somatic tissue and organ damage caused by either viral attack ^[12]^ or inflammation ^[10]^. These clinical assumptions lack molecular evidence, potentially leading to biased assessments. Moreover, these explanations failed to consider the metabolic role of LDH to balance excess lactate during hypoxia. Here we have systematically explored the proteome and metabolome of sera from COVID-19 patients with low and high serum LDH, and identified the specific host responses, which shed light on the pathogenesis and convalesce of COVID-19.

## MATERIALS AND METHODS

### Patient Information

We collected and curated the electronic medical records of patient information in Taizhou Hospital of Zhejiang Province, between January 17 and February 20, 2020. 212 patients met the criteria of suspected COVID-19, of which 145 patients were confirmed as COVID-19, based on the Government’s Diagnosis and Treatment Guideline (5th version) ^[14]^.144 patients COVID-19 patients were included in this study after excluding one patient who had incomplete laboratory data. According to the admission period, we grouped them into two cohorts. Patients admitted to Taizhou Public Health Medical Center, Taizhou Hospital between January 17 and February 4, 2020 were included in the cohort 1 (n = 115). The cohort 2 (n = 29) contained COVID-19 patients admitted from February 4 to February 20, 2020. The end of the follow-up date was March 1, 2020. Disease severity was accessed according to the abovementioned guideline. We classified COVID-19 patients into two groups (severe and non-severe): the severe group included severe and critical patients, and the non-severe group included mild and typical patients. Briefly, those who had shortness of breath with respiratory rate ≥ 30 breaths/min, a ratio of arterial blood oxygen partial pressure to oxygen concentration ≤ 300 mmHg or saturation of oxygen ≤ 93% when resting were defined as severe patients. The other COVID-19 patients were grouped as non-severe patients. 125 healthy individuals were enrolled as controls.

This study was conducted in accordance with the Helsinki Declaration and was approved by the institutional medical ethics review boards of Taizhou Hospital of Zhejiang Province and Westlake University (Approval ID: 20210119GTN001).

### Laboratory tests

Samples were taken throughout disease course from patient admission to discharge. More details are described in Supplementary Table 1. For laboratory tests, 404 serial blood samples from 144 patients were collected and centrifuged at 1500 g for 10 min at room temperature.

For serum of COVID-19 patients, seven biochemical indicators were tested, namely lactate dehydrogenase (LDH), aspartate aminotransferase (AST), alanine aminotransferase (ALT), total bilirubin, total protein, creatinine, and creatine kinase (CK), with a Chemistry Analyzer (Beckman Coulter, AU5821). The serum LDH was measured for healthy controls.

For LDH isoform analyses, 32 serum samples from 32 COVID-19 patients (11 non-severe and 21 severe) were sent for electrophoresis with agarose gel electrophoretic analyzer (SEBIA, HYDRASYS 2). ISO-LDH substrate and blocking buffer were afterwards added to incubate for 20 min, respectively. The gel was scanned with the same analyzer.

### Proteomic and metabolomic data set

The proteomic and metabolomic data were extracted from our previous publication ^[14]^. Briefly, serum samples from COVID-19 patients were kept at 56°C for 30 min to inactivate potential SARS-CoV-2. For proteomic experiments, inactivated serum samples were processed into peptides, labeled with TMTpro 16plex chemical tags, fractionated to 40 aliquots, and analyzed by LC-MS/MS. The proteomics data were analyzed with Proteome Discoverer (Version 2.4.1.15, Thermo Fisher Scientific), with the default parameter and a protein database composed of the Homo sapiens fasta database (07 Jan 2020, UniProtKB), containing 20,412 reviewed protein sequences, and the SARS-CoV-2 virus fasta (version NC_045512.2, NCBI). Targeted false discovery rate (FDR) for peptide-spectrum match was set to 1% (strict) and 5% (relaxed). Normalization was performed against the total peptide intensity. 894 proteins were quantified altogether. For metabolomic experiments, inactivated serum samples were processed to collect metabolites, and divided into 4 fractions for 4 different modes of LC-MS/MS data acquisition, leading to characterization of 941 metabolites. The median coefficient of variance (CV) for proteomic and metabolomic data were 10% and 5%, respectively, determined by pooled control samples in each batch, as described previously ^[14]^. The proteomics and metabolomics data could be referenced from the previously publication in ProteomeXchange Consortium (https://www.iprox.org/). Project ID: IPX0002106000 and IPX0002171000.

### Statistical analysis

Statistical clinical data analyses were performed using SPSS software (version 22.0). Continuous variables were presented as median and interquartile range (IQR) values, while categorical variables were shown as frequency and percentage. An independent t-test was used for continuous variables when the data were normally distributed; otherwise, the Mann-Whitney test was used. Chi-square test or Fisher’s exact test was used for categorical variables. Receiver operating characteristic (ROC) analysis was used for the selection of the best intercept point. Prediction of disease progression was obtained using the Cox proportional hazards model. Statistical proteomic data analysis was performed using R (version 3.6.3). Missing values in the proteomic data matrix were assigned as 0.01 unless otherwise mentioned. *P* values ≤ 0.05 were considered statistically significant unless otherwise mentioned. Differential protein expression was based on the cutoff: *P* values ≤ 0.05, |log_2_FC| > 0.25. Plotting was performed with R (version 3.6.3).

## RESULTS AND DISCUSSION

### Demographic, clinical, and laboratory characteristics

A total of 144 COVID-19 patients were enrolled in the study. Detailed demographic, clinical, and laboratory characteristics of these patients on admission were provided in Table 1. The median age was 47 years old, and 53.5% of them were male. The severe patients account for 25% (36/144) of the group, were 10 years older than the non-severe patients (55 vs. 45, *p* < 0.001), and were more likely to have fever symptoms on admission (*p* = 0.001). Severe patients received higher percentages of treatment in oxygen inhalation (p < 0.001), antibiotics (p = 0.024), glucocorticoid (p < 0.001), and intravenous gamma immunoglobulin (p < 0.001) than the non-severe patients. These medications may have an impact on the patient blood metabolism and may alter the COVID-19 microenvironment, which unfortunately could not be rigorously examined in the current study due to the small sample size, awaiting future investigations. The patients did not exhibit significant difference between severe and non-severe groups in terms of gender unmentioned symptoms on admission, nor other medical treatments (*p* > 0.05) as listed in Table 1.

**Table 1.** Clinical characteristics and Laboratorial indexes of COVID-19 patients.

| Characteristics | Total (N=144) | Non-severe<br>(N=108) | Severe (N=36) | *p* value |
| --- | --- | --- | --- | --- |
| **Age, years** |  |  |  |  |
| median (IQR) | 47 (38-56) | 45 (37-54) | 55 (48-65) | 0.000 |
| Sex, Male | 77 (53.5) | 57 (52.8) | 20(55.6) | 0.772 |
| **Symptoms on Admission** |  |  |  |  |
| Fever, % | 104 (72.2) | 70(64.8) | 34 (94.4) | 0.001 |
| Pharyngalgia, % | 17 (11.8) | 15 (13.9) | 2 (5.6) | 0.180 |
| Cough, % | 65 (45.1) | 47 (43.5) | 18 (50.0) | 0.499 |
| Expectoration, % | 26 (18.1) | 19 (17.6) | 7 (19.4) | 0.802 |
| Tiredness, % | 16 (11.1) | 10 (9.3) | 6 (16.7) | 0.221 |
| Headache, % | 16 (11.1) | 9 (8.3) | 7 (19.4) | 0.066 |
| **Medical Treatment** |  |  |  |  |
| Oxygen inhalation, % | 97 (67.4) | 63 (58.3) | 34 (94.4) | 0.000 |
| Antibiotics, % | 26 (18.1) | 15 (13.9) | 11 (30.6) | 0.024 |
| Antivirus, % | 144 (100) | 108 (100) | 36 (100) | 1.000 |
| Glucocorticoid, % | 37 (25.7) | 13 (12.0) | 24 (66.7) | 0.000 |
| Gamma Immunoglobulin, % | 33 (22.9) | 7 (6.5) | 26 (72.2) | 0.000 |
| **Laboratorial Indexes** |  |  |  |  |
| LDH (ULN) <sup>a</sup> |  |  |  | 0.000 |
| Low or normal | 115(79.9) | 100 (92.6) | 15 (41.7) |  |
| High | 29 (20.1) | 8 (7.4) | 21 (58.3) |  |
| ALT (ULN) <sup>b</sup> |  |  |  | 0.023 |
| Low or normal | 130 (90.3) | 101 (93.5) | 29 (80.6) |  |
| High | 14 (9.7) | 7 (6.5) | 7 (19.4) |  |
| AST (ULN) <sup>c</sup> |  |  |  | 0.000 |
| Low or normal | 125(86.8) | 101 (93.5) | 24 (66.7) |  |
| High | 19 (13.2) | 7 (6.5) | 12 (33.3) |  |
| Total bilirubin (ULN) <sup>d</sup> |  |  |  | 0.422 |
| Low or normal | 122(84.7) | 93(86.1) | 29 (80.6) |  |
| High | 22 (15.3) | 15 (13.9) | 7 (19.4) |  |
| Total protein (LLN) <sup>e</sup> |  |  |  | 0.477 |
| High or normal | 114 (79.2) | 87 (80.6) | 27 (75.0) |  |
| Low | 30 (20.8) | 21 (19.4) | 9 (25.0) |  |
| Urea (ULN) <sup>f</sup> |  |  |  | 0.004 |
| Low or normal | 137 (95.1) | 106 (98.1) | 31 (86.1) |  |
| High | 7 (4.9) | 2 (1.9) | 5 (13.9) |  |
| Creatinine (ULN) <sup>g</sup> |  |  |  | 0.029 |
| Low or normal | 135 (93.8) | 104 (96.3) | 31 (86.1) |  |
| High | 9 (6.3) | 4 (3.7) | 5 (13.9) |  |
| Creatine kinase (ULN) <sup>h</sup> |  |  |  | 0.000 |
| Low or normal | 123 (85.4) | 99 (91.7) | 24 (66.7) |  |
| High | 21 (14.6) | 9 (8.3) | 12 (33.3) |  |

Abbreviations: LDH, Lactate dehydrogenase; AST, aspartate aminotransferase; ALT, Alanine aminotransferase.
<sup>a</sup> ULN of LDH for male and female are 285 and 227 U/L, respectively.
<sup>b</sup> ULN of ALT for male and female are 50 and 40 U/L, respectively.
<sup>c</sup> ULN of AST for male and female are 40 and 35 U/L, respectively.
<sup>d</sup> ULN of total bilirubin is 20.5 µmol/L.
<sup>e</sup> LLN of total protein is 65 g/L.
<sup>f</sup> ULN of urea for male and female are 8 and 8.8 U/L, respectively.
<sup>g</sup> ULN of creatinine for male and female are 104 and 84 µmol/L, respectively.
<sup>h</sup> ULN of CK for male and female are 172 and 140 U/L, respectively.

Based on laboratory test results, a higher percentage of severe patients have elevated levels of serum LDH that are above the upper limit of normal (ULN) value than non-severe patients (58.3% vs. 7.4%, *p* < 0.001). Likewise, more severe patients also showed higher level of alanine aminotransferase (ALT, *p* = 0.023), aspartate aminotransferase (AST, *p* < 0.001), urea (*p* = 0.004), creatinine (*p* = 0.029), and creatine kinase (CK, *p* < 0.001). We then compared their temporal changes at 7-day intervals (Supplementary Table 1). LDH, CK, and creatinine showed continuous decrease in the sera of severe patients, while only serum LDH showed a continuous decrease in non-severe patients.

A Cox regression model was applied to evaluate the prognostic value of these indicators (Supplementary Table 2). In the Cohort 1, LDH is the only indicator with discrimination ability (p < 0.001) in the multivariate analysis, although LDH, AST, and CK were determined in the univariate analysis with statistical significance (p < 0.001). We further validated the value of serum LDH in Cohort 2 and the data showed significant discrimination (*p* = 0.032), suggesting that serum LDH could be a useful prognostic biomarker.

### Temporal characteristics of Serum LDH in COVID-19 patients

We then assessed the effect of age and sex on the temporal serum LDH expression. COVID-19 patients older than 60 years showed a higher serum LDH expression throughout the disease course than that in younger patients (Figure 1A), probably due to ageing and underlying diseases. Males expressed higher serum LDH levels than that from females only during the initial hospitalization stage (within the first week) (Figure 1B). Serum LDH levels became comparable between COVID-19 patients and healthy controls (median values: 177 U/L above 60 years old and 152 U/L below 60 years old; 154 U/L for males and 158.5 U/L for females) after three weeks.

**Figure 1.**
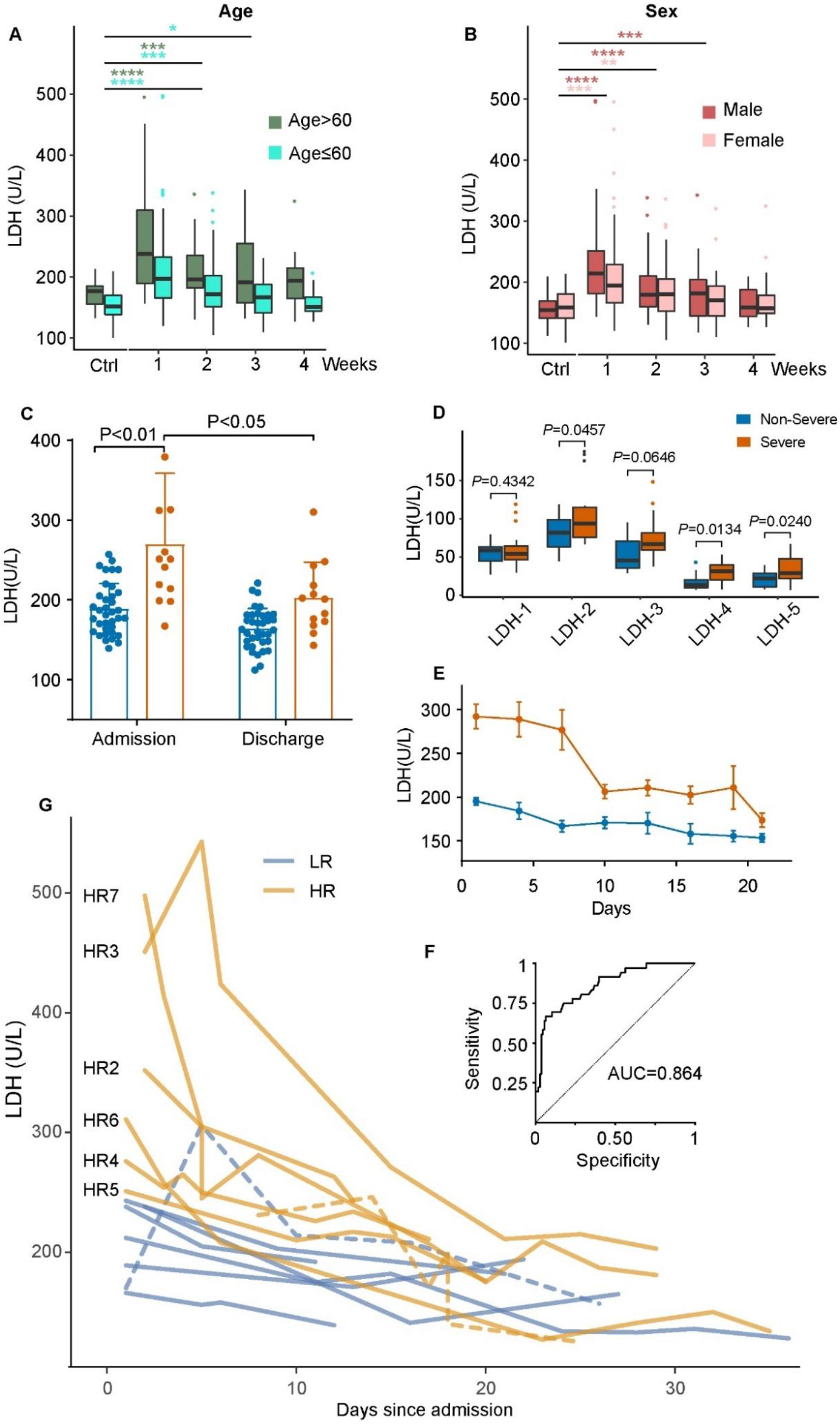
Serum LDH expression levels in the study cohort. A-B) Serum LDH expression levels in different time courses grouped by age and sex. *, 0.01<p≤0.05; **, 0.005<p≤0.01; ***, 0.001<p≤0.005; ****, p≤0.001. C) Serum LDH expression levels of severe and non-severe patients during the admission and discharge stage. D) Expression of serum LDH isoforms during the admission stage. E) Serum LDH expression levels of severe and non-severe patients in time courses at the 3-day interval. F) Receiver operating characteristic (ROC) of the study cohort when setting LDH expression level cutoff as 247 U/L. G) Individual inspection of 14 patients’ LDH expression dynamics. Yellow dash line, Patient HR1; Purple dash line, Patient LR4.

We compared the median values of serum LDH levels from the initial admission stage (1-3 days after admission) and discharge stage (1-3 days before discharge), in a sub-cohort of recovered patients that were discharged by the end of follow-up. (total: n=49; severe: n=13; non-severe: n=36) (Figure 1C). Serum LDH levels were significantly higher in the severe patient group, particularly on admission (*p* < 0.01).

We analyzed serum LDH isoform expression from 32 patients upon admission (severe: n=22; non-severe: n=11) (Figure 1D) and found that LDH-2, LDH-4 and LDH-5 were significantly higher in the sera of severe COVID-19 patients. LDH-1 in contrast was not dysregulated. LDH-4 and LDH-5 mainly contribute to pyruvate fermentation under hypoxic conditions, while LDH-1 favors the opposite direction of the reversible reaction ^[15]^. These data suggest anaerobic glycolysis metabolism in severe patients.

To better understand the temporal dynamics of serum LDH expression over the disease course, we next monitored the serum LDH level from day 1 on admission till day 21 at a 3-day interval (Figure 1E). As to the non-severe patient group, the serum LDH level was slightly higher on admission while declining slowly over the hospitalization period. The serum LDH levels in the severe group comparatively were significantly higher upon admission, with a prominent variance range. They dropped significantly from 3rd to 9th day as the patients were taking medical care and by the 21st day fell below the initial serum LDH levels in the non-severe group.

### Classification of low- and high-risk patients based on serum LDH levels

To establish a cohort-specific serum LDH expression threshold as a risk indicator, we took patient severity (severe vs. non-severe) as the dichotomous variable and conducted the receiver operating characteristic (ROC) analysis on the serum LDH expression levels on admission. The area under curve (AUC) was 0.864 (Figure 1F), confirming the discriminative power of serum LDH. The serum LDH level corresponding to the maximum Youden index was determined as 247 U/L, within the threshold to determine serum LDH abnormality from past reports (240-253.2 U/L) ^[13]^.

Fourteen characteristic patients within the cohort were selected and divided into two groups for closer inspection (Figure 1H). The low-risk (LR) group consists of seven non-severe patients with serum LDH expressions below 247 U/L since admission except for one exceptional detection (LR4, 5^th^ day, 306 U/L). The high-risk (HR) group are composed of six severe patients with on admission serum LDH levels above 247 U/L, and one severe patient with serum LDH levels below the threshold throughout hospitalization (HR1). We attribute this to the relatively late first sampling timepoint (8th day), given that serum LDH levels from all the patients started at a relatively high level and declined over time. Patient HR3 and patient HR7 in particular had exceptionally high serum LDH levels (> 450 U/L) upon admission but dropped dramatically within 10 days. We inspected the detailed medical records of the 14 patients (Table 2). COVID-19 patients with comorbidities including hypertension, chronic HBV infection, and diabetes tend to be severe patients in the HR group, in consistent with the literature ^[16,17]^.

**Table 2.**
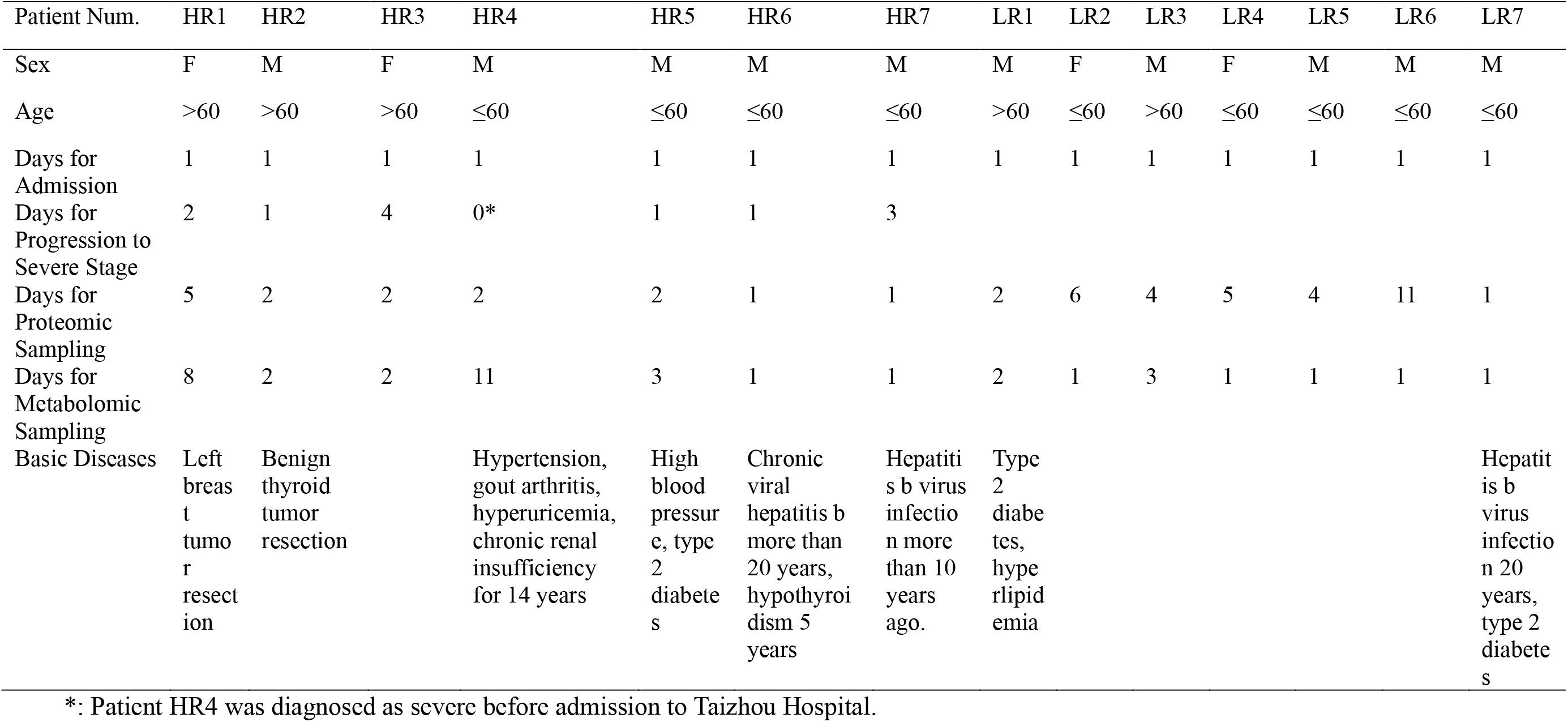
Medical records of temporal change associated severe patients.

| Patient Num. | HR1 | HR2 | HR3 | HR4 | HR5 | HR6 | HR7 | LR1 | LR2 | LR3 | LR4 | LR5 | LR6 | LR7 |
| --- | --- | --- | --- | --- | --- | --- | --- | --- | --- | --- | --- | --- | --- | --- |
| Sex | F | M | F | M | M | M | M | M | F | M | F | M | M | M |
| Age | >60 | >60 | >60 | ≤60 | ≤60 | ≤60 | ≤60 | >60 | ≤60 | >60 | ≤60 | ≤60 | ≤60 | ≤60 |
| Days for Admission | 1 | 1 | 1 | 1 | 1 | 1 | 1 | 1 | 1 | 1 | 1 | 1 | 1 | 1 |
| Days for Progression to Severe Stage | 2 | 1 | 4 | 0* | 1 | 1 | 3 |  |  |  |  |  |  |  |
| Days for Proteomic Sampling | 5 | 2 | 2 | 2 | 2 | 1 | 1 | 2 | 6 | 4 | 5 | 4 | 11 | 1 |
| Days for Metabolomic Sampling | 8 | 2 | 2 | 11 | 3 | 1 | 1 | 2 | 1 | 3 | 1 | 1 | 1 | 1 |
| Basic Diseases | Left breast tumor resection | Benign thyroid tumor resection |  | Hypertension, gout arthritis, hyperuricemia, chronic renal insufficiency for 14 years | High blood pressure, type 2 diabetes | Chronic viral hepatitis b more than 20 years, hypothyroidism 5 years | Hepatitis b virus infection more than 10 years ago. | Type 2 diabetes, hyperlipidemia |  |  |  |  |  | Hepatitis b virus infection 20 years, type 2 diabetes |

\*: Patient HR4 was diagnosed as severe before admission to Taizhou Hospital.

### Quantitative proteomics and metabolomics uncover dysregulated molecules associated with elevated serum LDH

The serum proteomic and metabolomic datasets of the HR and LR group patients were extracted from a collateral project ^[14]^. 78.6% (11/14) of the patient sera were sampled during the first week on admission (Table 2), herein representing the stage when serum LDH levels exhibited sharp difference between the LR and HR groups. For the proteomic dataset, the Student’s *t*-test highlighted 34 proteins as differentially expressed (*p* < 0.05) between HR and LR groups (Figure 2A, upper panel), 26 of which were up-regulated. Pathway enrichment analysis using Metascape ^[18]^ showed these proteins conduct three major immune-related activities including acute inflammatory responses (GO:0002526, *p* < 0.001), platelet degranulation (GO:0002576, *p* < 0.001) and regulation of complement cascade (R-HSA-977606, *p* < 0.001) (Figure 2A and Supplementary Fig. 2A). Additional analyses using Ingenuity Pathway Analysis (IPA) nominated acute phase response signaling as the most activated immune-related pathway (Supplementary Fig. 2C) in HR patients. These findings were in consistence with the prominent immune behaviors (Supplementary Fig. 1B) as we have previously reported in COVID-19 patients with different severity ^[14]^. Moreover, blood coagulation (GO: 007596, *p* < 0.001) was significantly enriched (Supplementary Fig. 2A and 2C). This pathway has been reported to be altered in COVID-19, and associated with interleukin-6 (IL-6))^[19]^. Our data showed upregulation of acute phase proteins (SAA1, ORM1, AGT, and SERPINA3), complement subunits (C9, C6, and CFI), and LDH subtypes (LDHA and LDHB) in the HR group (Figure 2B). Thirteen of the differentiated proteins were mapped into a network wherein key regulators were focused (Figure 2C). Within them, pro-inflammatory cytokine IL-6 has been widely recognized as a risk factor for COVID-19 ^[19-23]^ and clinically observed to be positively correlated with serum LDH levels ^[24]^. IL-6 can activate TP53, which facilitates cell apoptosis and could enhance LDHA expression in blood. Multiple COVID-19 studies involving IL-6 agree with our profiling ^[19,25]^. This network also includes CCAAT/enhancer-binding protein beta (CEBPB) which mediates immune and inflammatory responses ^[26]^, and sterol regulatory element binding transcription factor 1 (SREBF1) which regulates lipid metabolisms that has been reported dysregulated in severe COVID-19 patients ^[14]^. Taken together, the proteomic difference between LR and HR patients reflected different host responses between the non-severe and severe COVID-19 patients.

**Figure 2.**
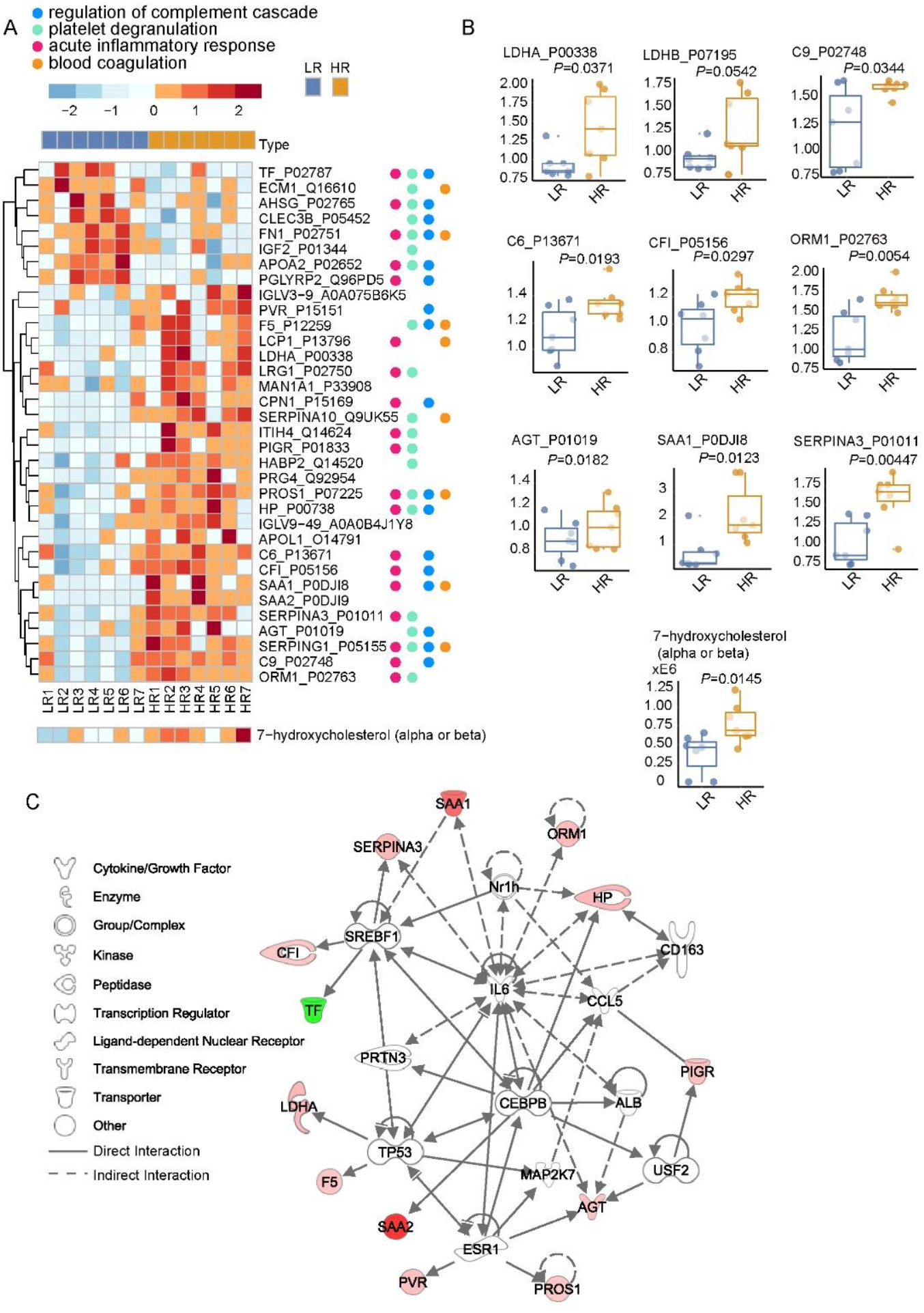
Molecular differences between low- and high-risk patients. A) Heatmap of 34 differentially expressed proteins and two differentially expressed metabolites. LR, low-risk patients. HR, high-risk patients. B) Boxplots of nine selected differentially expressed proteins and two selected differentially expressed metabolites. C) Protein network including 12 selected differentially expressed proteins.

Of the 34 differentially expressed metabolites listed in Supplementary Fig. 3A, 88.2% (30/34) were upregulated, and 52.9% (18/34) were lipids. Within them, 7-hydroxycholesterol (beta) mediates oxidative stress and induces cell apoptosis. It was elevated during hepatitis B virus (HBV) and hepatitis C virus (HCV) infections ^[27]^ (Fig 2A and Fig 2B, lower panel). Further calibration and absolute quantification of these lipids would enable in-depth characterization of lipids and their variants. The protein-metabolite joint network generated using IPA (Supplementary Fig. 3B) proposed two upstream molecules to regulate LDHA expression, including hypoxia-inducible factor 1 (HIF1) that mediates hypoxic and inflammatory microenvironments ^[28]^ and fibronectin 1 (FN1) that involves COVID-19 lung fibrosis ^[29]^. The dysregulated metabolites further consolidate disturbed host responses in association with serum LDH increase uncovered by the proteomic data.

### Protein and metabolite change in patients with exceptionally high serum LDH

Next, we narrowed our focus to the two patients with exceptionally high serum LDH levels on admission (HR3 and HR7, HR outliers), and compared their proteomic patterns with the other HR patients (Figure 3A). 38 proteins including LDHA and LDHB were differentially expressed (*p* < 0.05, Figure 3A). Metascape pathway enrichment nominated 23 proteins associated with immune system process (GO:0002376, *p* < 0.001) and 21 proteins associated with metabolic process (GO:0008152, *p* < 0.001) (Supplementary Fig 4B). The top enriched immune-related pathways are the activation of immune response (GO:0002253, *p* < 0.001) and humoral immunity response (GO:0006959, *p* < 0.001), while the top enriched metabolic pathway relates to cofactors (R-HSA-89 78934, *p* < 0.001) (Supplementary Fig. 4A). IPA analysis detailed the top metabolic functions as protein ubiquitination pathway and pyruvate fermentation to lactate (Supplementary Fig 4C). Especially, the HIF1α signaling pathway showed a drastic activation in the HR (outliers) group. As for characteristic proteins in HR (outliers), our data uncovered a dysregulated protein group which includes five up-regulated proteasome subunits, namely PSMA3, PSMA4, PSMA5, PSMB1, and PSMB3 (Figure 3B), possibly due to cell apoptosis ^[30]^ from organ/tissue damage. They are the mediators of protein ubiquitination and associate with NF-κB signaling (GO:0038061, *p* < 0.001). Six of the seven differentially expressed immunoglobulin residues (IGHV3-43, IGHV3-30-5, IGKV1-5, IGLV1-36, IGKV1-17, and IGKV2-24) (Supplementary Fig. 5A) were up-regulated in HR (outliers), suggesting that humoral immunity at the point of detection was suppressed or not activated. For the other up-regulated proteins in HR (outliers) (Figure 3B), CES1 is a hepatic protein and its release in blood suggests liver injuries. Protein disulfide-isomerase (P4HB) was reported to up-rise in response to hypoxia ^[31]^. GAPDH could enhance HIF activity ^[32]^ via NF-κB induction activated in hypoxia ^[33]^, which contributes to heat shock protein 90-alpha (HSP90AA1) upregulation ^[34]^ to form protein complexes with Hif1α. Dipeptidyl peptidase 4 (DPP4), also known as CD26, has been nominated as a potential critical marker in infection susceptibility ^[35]^, and its inhibition has been proposed to reduce COVID-19 patient severity ^[36]^. DPP4 is also a downstream factor to mark HIF pathway induction ^[37]^. Especially, the down-regulated protein in HR (outliers) includes CPB2 as a basic carboxypeptidase that suppresses complement system-mediated inflammation ^[38]^. Its deficiency could lead to accelerated acute lung injuries ^[39]^. 84.6% (11/13) of the dysregulated metabolites were lipids (Supplementary Fig. 5B), suggesting disturbed lipid metabolism accompanied with serum LDH changes. The network analysis (Supplementary Fig. 5C) further proposed LDH elevation to be associated with HSP90AA1 and proteasomes.

**Figure 3.**
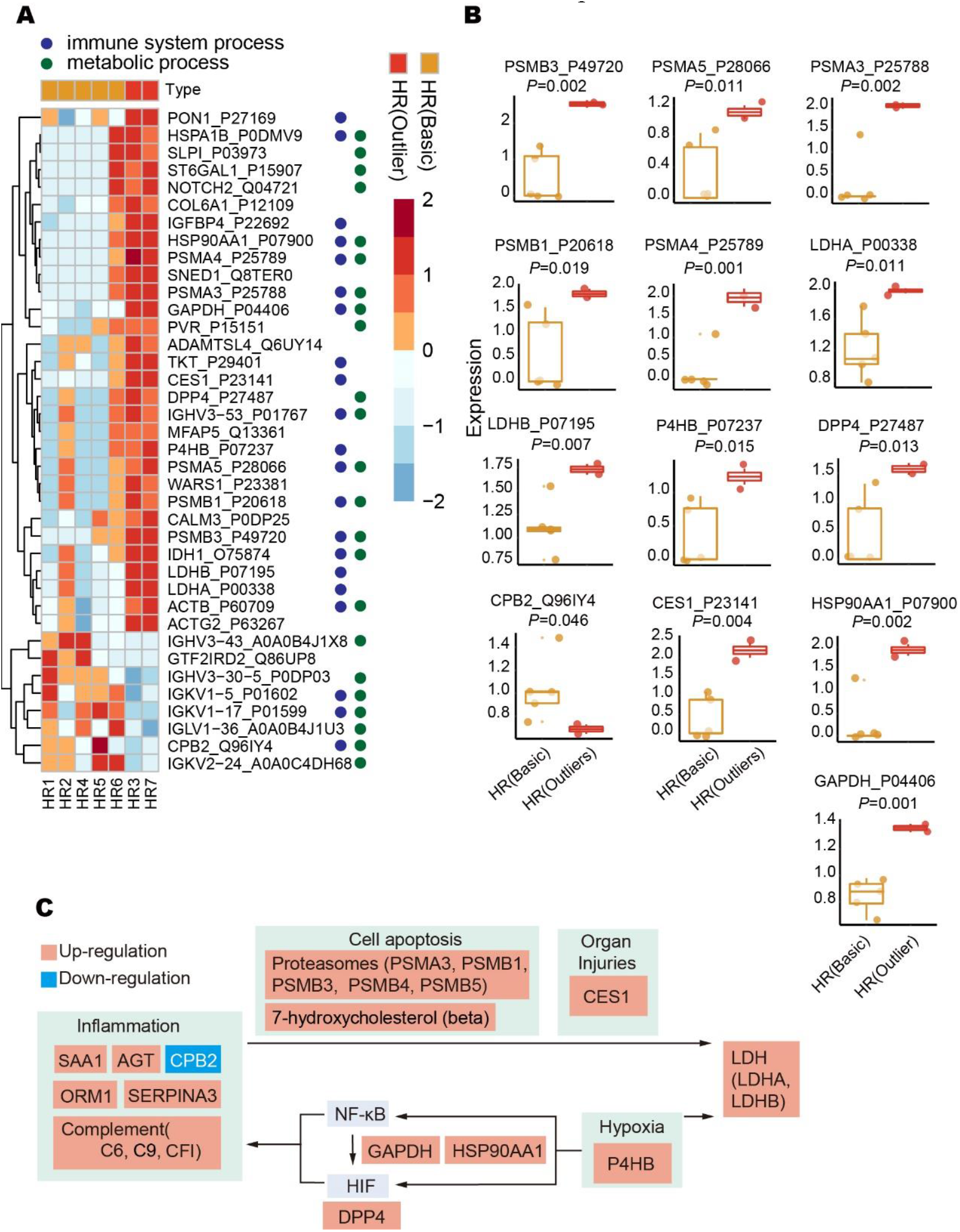
Molecular differences between HR (basic) and HR (outliers) patients. A) Heatmap of 38 differentially expressed proteins. LR, low-risk patients. HR, high-risk patients. B) Boxplots of 13 selected differentially expressed proteins. C) Proposed mechanism for serum LDH elevation in COVID-19 patients.

### Serum LDH elevation might be driven by tissue injuries and hypoxia

Taking together all the perturbed molecules as highlighted above (Figure 2B and 3B), we propose a putative working model for the serum LDH elevation in COVID-19 patients (Figure 3C). On the one hand, the inflammation processes triggered by the host immune system induce apoptosis of the infected cells, leading to the release of intracellular LDH into the blood. In high-risk cases, these immune activities result in over-reactive inflammation processes (like “cytokine storm”) ^[40]^, thereby releasing higher levels of serum LDH from multiple organs/tissues ^[10]^. On the other hand, oxygen homeostasis was disturbed in severe COVID-19 patients ^[12]^. Hypoxia reactions occur to accumulate lactate via glycolysis. LDH can balance lactate secretion via pyruvate fermentation and a series of metabolic regulation (Supplementary Fig. 4A and 4C) to maintain cellular homeostasis ^[41]^. The activated NF-κB and HIF pathways in hypoxia conditions could also induce inflammatory responses ^[42]^.

## CONCLUDING REMARKS

Here we systematically investigated serum LDH elevation in COVID-19 patients. We thoroughly inspected both clinical and molecular profiles of 144 COVID-19 patients. We confirmed serum LDH as the best independent risk indicator, and further optimized a threshold of 247 U/L for stratifying COVID-19 patients based on the serum LDH level. Our data showed that the serum LDH declined thereafter, patients with serum LDH levels higher than the threshold on admission are prone to severe conditions, hence determined as high-risk (HR) patients, and those lower than the threshold as low-risk (LR) patients.

Proteomic differences between LR and HR groups exposed a list of dysregulated host responses. Among them, acute inflammatory responses, platelet degranulation and complement cascade have been reported in previous studies comparing severe and non-severe COVID-19 patients ^[14,43]^. Blood coagulation has been highlighted in another report that compares COVID-19 patients with high and low IL-6 levels ^[44]^. Immune behaviors including activation of immune response and humoral immune response were further enriched during the intra-comparison within HR patients, suggesting that the immune behaviors are closely related to serum LDH expression. Proteomic profiling also highlighted a list of hypoxia related proteins and functions, including P4HB, DPP4, GAPDH, HSP90AA1, NF-κB and HIF signaling, suggesting that hypoxia might have contributed to elevated LDH. The metabolomic profiling complements findings on the proteomic level and further emphasizes dysregulated lipid metabolism. Taken together, we propose that elevation of serum LDH might attribute to inflammation-related tissue injuries and hypoxia-related metabolism.

This study is limited by several factors. Firstly, this is a single-center study with a relatively small patient cohort due to difficulties of sample collection, therefore subject to experimental bias. Also, the standard inactivation procedures for COVID-19 serum to minimize the risk of infection may have some impact on the characterized samples. Moreover, proteome data from only 14 individuals were acquired, from which only 2 patients were determined as high-risk group outliers for comparative analysis. We could not obtain the samples from different patients at the identical time points. And due to the small sample size, multiple testing wasn’t performed for molecular analyses, therefore the statistical power from the proteomic data has to be interpreted with caution. It is worth noting that serum LDH elevation is not specific to COVID-19 disease ^[45]^. Further studies should conduct clinical validation on larger cohorts, and compare the molecular differences including control patients with other diseases with similar symptoms. Targeted approaches would also be required to validate our findings for diagnostic purposes.

## Data Availability

All data referred to in the manuscript are available and can be accessed online.

http://www.iprox.org

## Data Availability

Data for this study are not currently publicly available. The Author intends to make the data open access in early 2021.

## ACKNOWLEDGMENTS

This work was supported by grants from National Key R&D Program of China (No. 2020YFE0202200), National Natural Science Foundation of China (81972492, 21904107, 81672086, 82072333), Zhejiang Provincial Natural Science Foundation for Distinguished Young Scholars (LR19C050001), Hangzhou Agriculture and Society Advancement Program (20190101A04), Zhejiang Provincial Natural Science Foundation of China (LQ19H100001), Zhejiang Medical and Health Science and Technology Plan (2021KY394), Westlake Education Foundation, and Tencent Foundation. We thank Westlake University Supercomputer Center for assistance in data storage and computation.

