## Supplementary for "Proteomic and Metabolomic Investigation of COVID-19 Patients with Elevated Serum Lactate Dehydrogenase"

**Supplementary Table 1** Dynamic changes of clinical laboratory test results in severe and non-severe COVID-19 patients

| Characteristics | Total COVID-19 patients (n=144) | | Sampling points | |
|---|---|---|---|---|
| | Severe (n=36) | non-Severe (n=108) | Severe | non-Severe |
| LDH, U/L | | | | |
| 1-7 day | 272 (218-312) | 187 (163-217) | 34 | 103 |
| 8-14 day | 216 (183-260) | 163 (139-189) | 28 | 55 |
| 15-21 day | 198 (180-214) | 157(132-185) | 18 | 32 |
| >21 day | 174 (146-207) | 151(145-165) | 15 | 20 |
| Total bilirubin, µmol/L | | | | |
| 1-7 day | 12.2 (7.8-17.6) | 12.3 (8.3-17.2) | 36 | 105 |
| 8-14 day | 14.2 (10.6-23.0) | 14.5 (11.4-17.0) | 36 | 87 |
| 15-21 day | 13.1 (7.9-18.0) | 11.8 (8.7-14.7) | 29 | 47 |
| >21 day | 9.9 (6.5-13.0) | 7.5 (6.5-11.6) | 17 | 32 |
| Urea, µmol/L | | | | |
| 1-7 day | 4.3 (3.5-5.7) | 3.9 (3.2-5.1) | 36 | 106 |
| 8-14 day | 6.0 (4.5-7.8) | 3.6 (3.0-4.2) | 34 | 87 |
| 15-21 day | 4.6 (3.6-6.1) | 3.7 (3.1-4.3) | 28 | 47 |
| >21 day | 4.1 (3.6-4.1) | 3.8 (3.0-4.5) | 21 | 28 |
| Total protein, g/L | | | | |
| 1-7 day | 68.7 (64.2-72.4) | 69.2 (66.0-74.0) | 36 | 107 |
| 8-14 day | 70.6 (65.9-75.3) | 67.0 (63.0-69.9) | 36 | 88 |
| 15-21 day | 65.7 (60.7-71.2) | 65.9 (64.3-70.5) | 30 | 47 |
| >21 day | 62.9 (58.2-67.0) | 68.9 (65.8-70.7) | 17 | 32 |
| AST, U/L | | | | |
| 1-7 day | 30.0 (25.3-50.0) | 22.0 (19.0-28.0) | 36 | 107 |
| 8-14 day | 23.5 (19.0-31.3) | 22.0 (17.0-26.0) | 38 | 87 |
| 15-21 day | 24.0 (18.0-36.5) | 21.0 (19.0-25.0) | 29 | 47 |
| >21 day | 22.0 (20.0-41.5) | 22.0 (20.0-26.0) | 17 | 31 |
| ALT, U/L | | | | |
| 1-7 day | 30.0 (19.0-39.0) | 19.0 (13.0-30.0) | 35 | 106 |
| 8-14 day | 35.5 (23.0-52.8) | 20.0 (15.0-34.0) | 40 | 83 |
| 15-21 day | 36.5 (20.8-55.5) | 23.0 (17.0-36.3) | 38 | 42 |
| >21 day | 27.0 (20.0-41.0) | 26.5 (19.5-35.5) | 27 | 42 |
| Creatine kinase, U/L | | | | |
| 1-7 day | 104.5 (67.5-209.3) | 55.5 (38.5-79.8) | 28 | 100 |
| 8-14 day | 31.0 (26.0-41.0) | 35.0 (27.0-50.0) | 29 | 67 |
| 15-21 day | 28.0 (22.0-39.0) | 43.0 (29.0-55.0) | 31 | 36 |
| >21 day | 26.0 (19.0-32.8) | 43.0 (32.0-56.0) | 20 | 15 |
| Creatinine, µmol/L | | | | |
| 1-7 day | 75.5 (66.0-93.0) | 71.5 (62.5-84.0) | 36 | 108 |
| 8-14 day | 75.0 (65.0-91.0) | 76.0 (63.0-85.0) | 36 | 87 |
| 15-21 day | 69.5 (60.5-92.8) | 71.0 (62.0-81.0) | 28 | 45 |
| >21 day | 67.0 (57.5-87.0) | 72.0 (62.0-79.3) | 17 | 30 |

**Supplementary Table 2** Cox regression model for univariate and multivariate analysis.

| Characteristics | Cohort 1 (n=115) | | | | Cohort 2 (n=29) | |
|---|---|---|---|---|---|---|
| | Univariate analysis | | Multivariate analysis | | Univariate analysis | |
| | HR (95%CI) | P value | HR (95%CI) | P value | HR (95%CI) | P value |
| Age | | | | | | |
| <60 | 1 | 0.019 | 1 | 0.155 | | |
| ⩾60 | 2.747 (1.183-6.380) | | 2.034(0.765-5.408) | | | |
| Sex | | | | | | |
| Male | 1 | 0.657 | | | | |
| Female | 0.834 (0.375-1.857) | | | | | |
| Fever | | | | | | |
| No | 1 | 0.022 | 1 | 0.061 | | |
| Yes | 10.336(1.398-76.429) | | 7.426 (0.915-57.395) | | | |
| Coexisting disorders | | | | | | |
| No | 1 | 0.197 | | | | |
| Yes | 1.676 (0.764-3.673) | | | | | |
| LDH (ULN) [a] | | | | | | |
| Low or normal | 1 | 0.000 | 1 | 0 | 1 | 0.032 |
| High | 11.396(5.037-25.786) | | 6.590 (2.415-17.981) | | 3.703 (1.120-12.244) | |
| ALT (ULN) [b] | | | | | | |
| Low or normal | 1 | 0.052 | | | | |
| High | 2.651 (0.993-7.076) | | | | | |
| AST (ULN) [c] | | | | | | |
| Low or normal | 1 | 0.000 | 1 | 0.598 | | |
| High | 5.368 (2.356-12.233） | | 1.329 (0.462-3.824) | | | |
| Total bilirubin (ULN) [d] | | | | | | |
| Low or normal | 1 | 0.183 | | | | |
| High | 1.867 (0.745-4.676) | | | | | |
| Total protein (LLN) [e] | | | | | | |
| High or normal | 1 | 0.746 | | | | |
| Lower | 0.838(0.288-2.442) | | | | | |

| | | | | |
|---|---|---|---|---|
| Urea (ULN) [f] | | | | |
| Low or normal | 1 | 0.025 | 1 | 0.125 |
| High | 3.982 (1.188-13.347) | | 4.836 (0.647-36.162) | |
| Creatinine (ULN) [g] | | | | |
| Low or normal | 1 | 0.048 | 1 | 0.306 |
| High | 2.939 (1.008-8.573) | | 0.389 (0.064-2.367) | |
| Creatine kinase (ULN) [h] | | | | |
| Low or normal | 1 | 0.000 | 1 | 0.545 |
| High | 5.214(2.357-11.534) | | 1.365(0.498-3.746) | |

### Supplementary Figures

**Supplementary Fig 1. Study design of COVID-19 patients with elevated serum lactate dehydrogenase.**

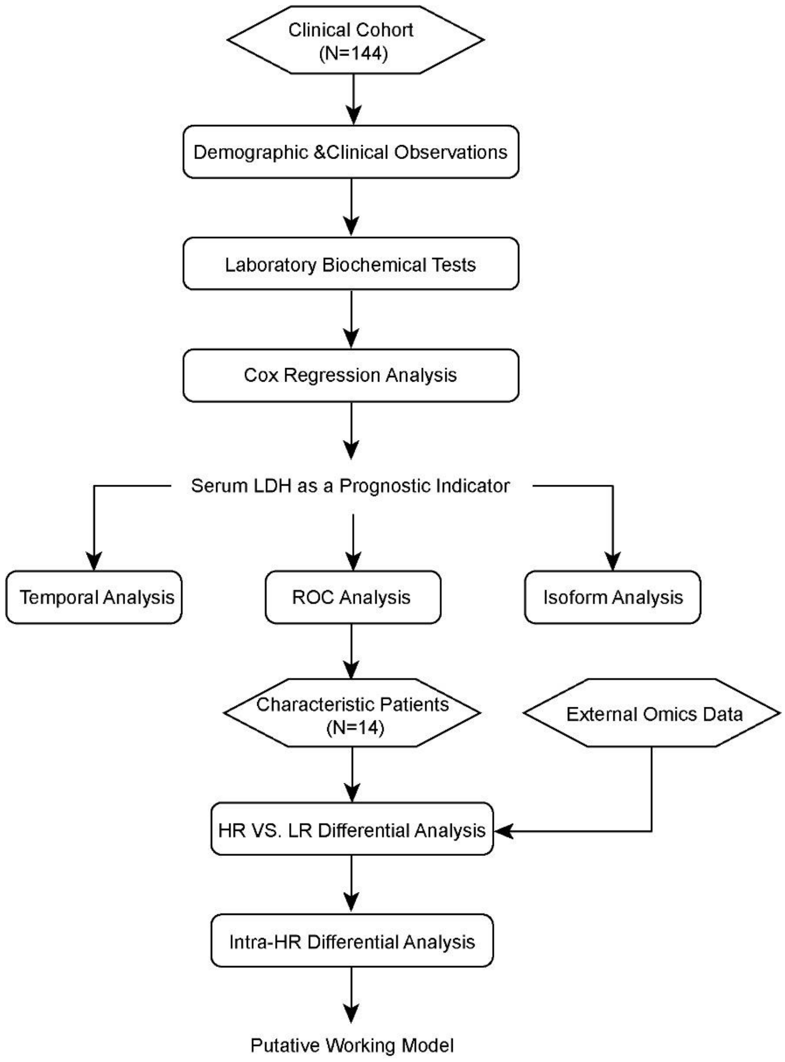

**Supplementary Fig 2. Pathway analyses of differentially expressed proteins between LR and HR patients.** A) Heatmap of Gene Ontology pathway enrichment using Metascape. B) Heatmap of Coronascape enrichment using Metascape. C) Barplot of Pathway enrichment using IPA.

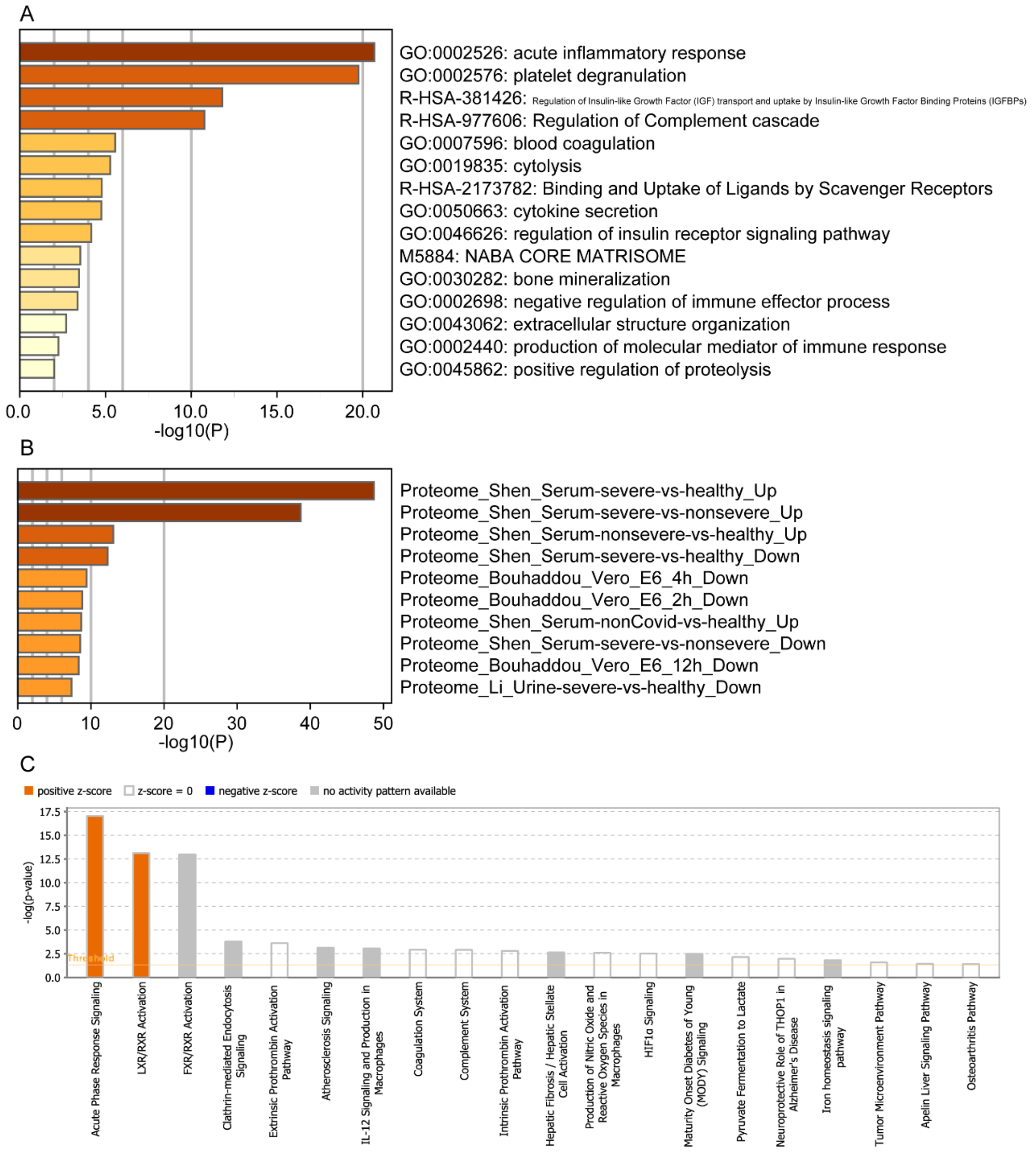

**Supplementary Fig3. Extended analyses of differentially expressed molecules between LR and HR patients.** A) Heatmap of differentially expressed metabolites. B) Protein-metabolic joint network analyses of differentially expressed molecules.

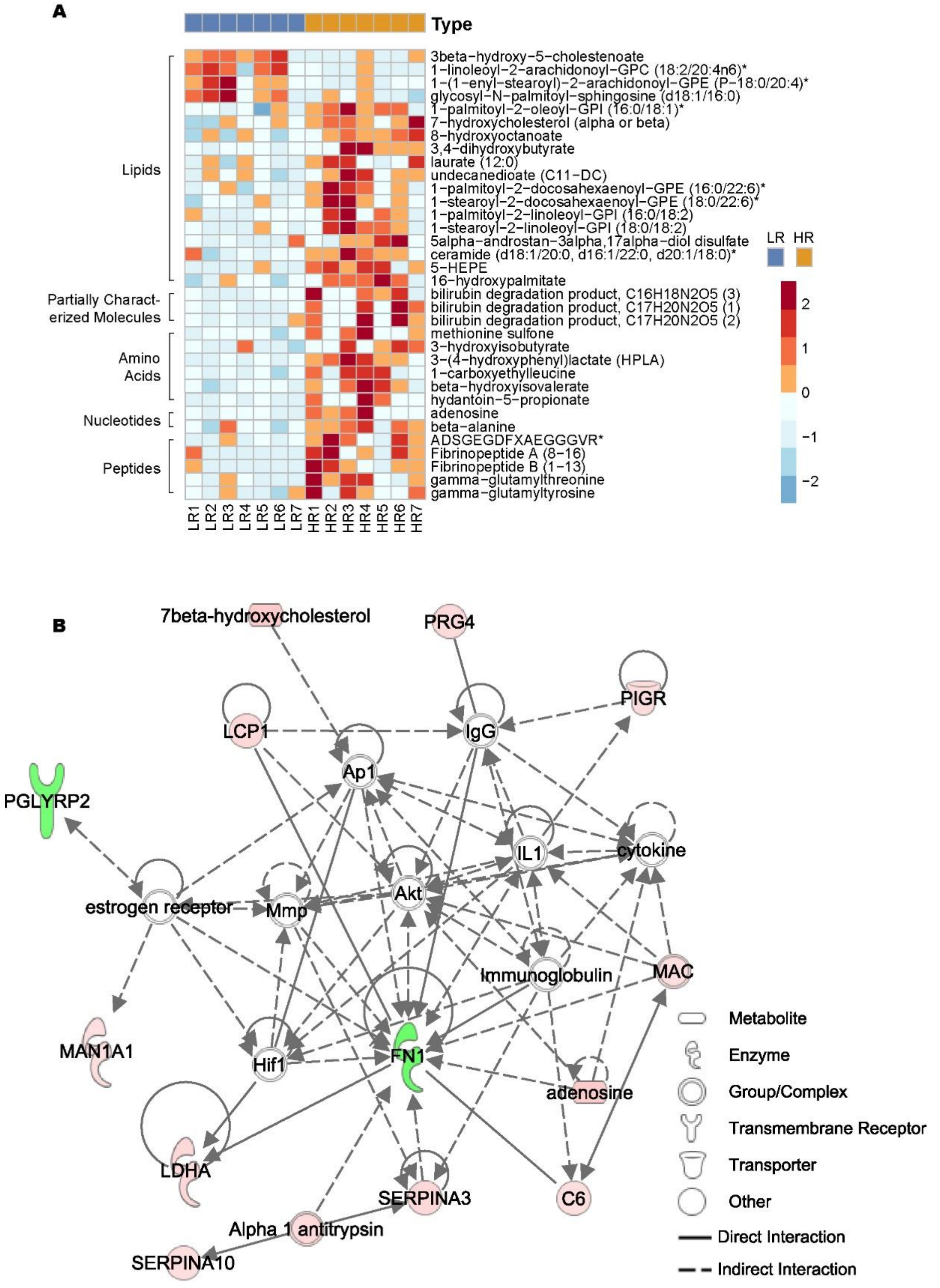

**Supplementary Fig 4. Pathway analyses of differentially expressed proteins between HR (Basic) and HR (Outliers) patients.** A) Heatmap of Gene Ontology pathway enrichment using Metascape. B) Heatmap of Gene Ontology (parent) pathway enrichment using Metascape. C) Barplot of Pathway enrichment using IPA

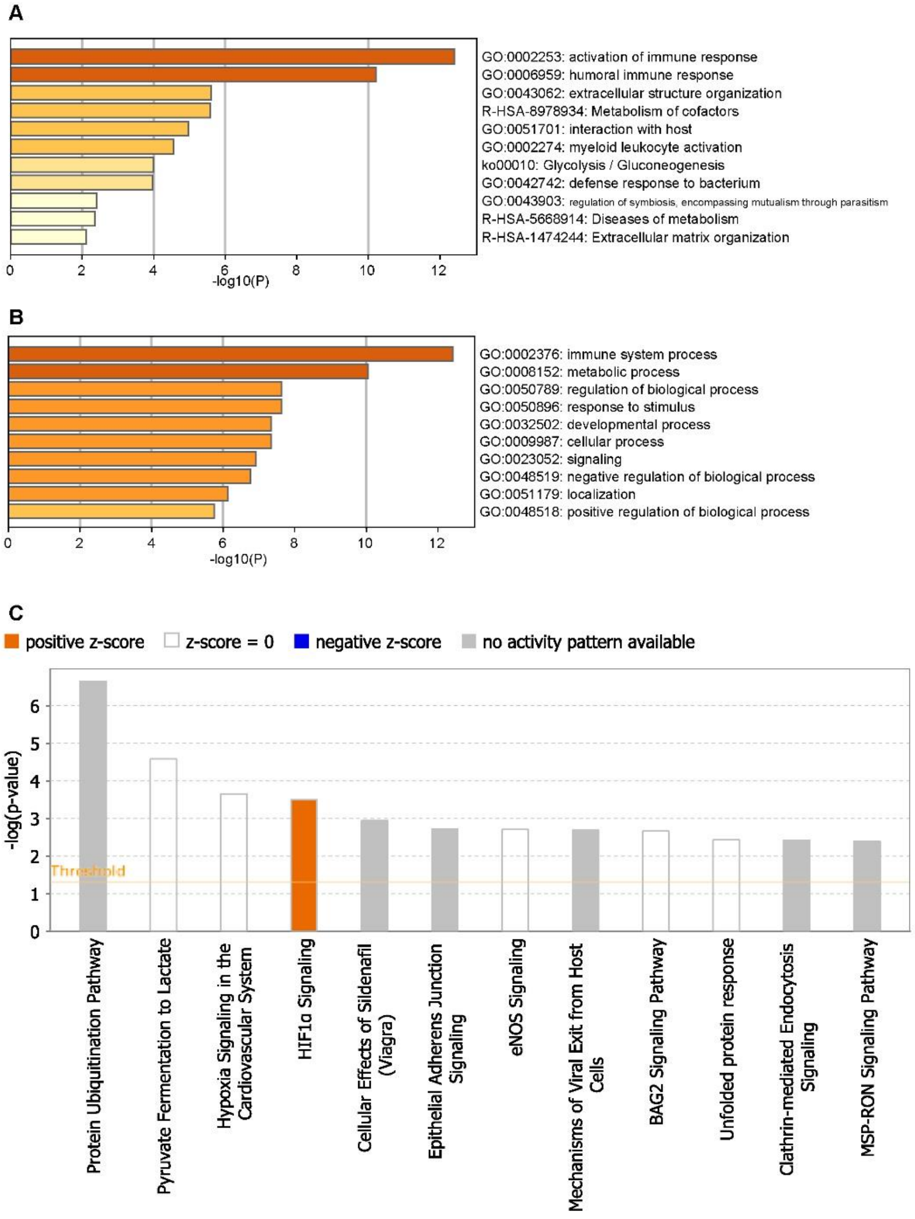


.

**Supplementary Fig5. Extended analyses of differentially expressed molecules between HR (Basic) and HR (Outliers) patients.** A) Heatmap of differentially expressed metabolites. B) Protein-metabolic joint network analyses of differentially expressed molecules.

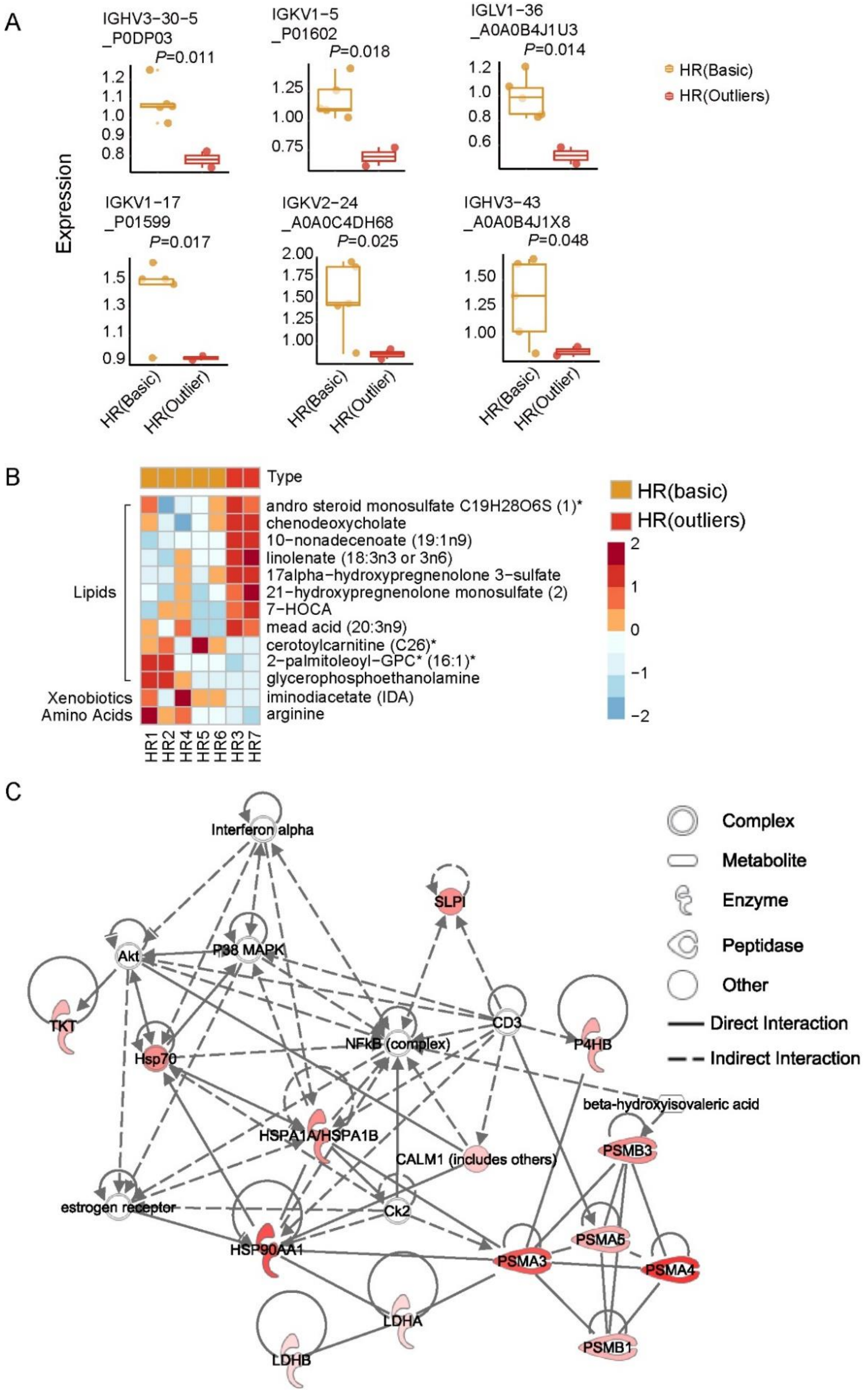
